# Medicaid Expansion and Inpatient Hospital Charges Among Women with Major Depressive Disorder

**DOI:** 10.1101/2025.10.07.25337532

**Authors:** Oluwasegun Akinyemi, Mojisola Fasokun, Fadeke Ogunyankin, Gabriella Kuffour, Samar K. Khalil, Ofure Omokhodion, Rachael Oyebade, Chioma Ekwunazu, Ayomide Ogunsakin, Miriam Michael, Guoyang Luo

**Affiliations:** The Clive O Callender Outcomes Research Center, Howard University College of Medicine, Washington DC, USA; Department of Internal Medicine, University Hospitals Cleveland Medical Center, Cleveland, Ohio, USA; Department of Research Data Science and Analytics, Cook Children’s Health Care System: Cook Children’s Medical Center, Fort Worth., Texas, USA; Department of Obstetrics and Gynecology, University of Central Florida, Orlando, FL, USA; Department of Family Medicine, University of Iowa Hospital and Clinics, Iowa City, IA, USA; Department of Internal Medicine, Howard University College of Medicine, Washington DC, USA; Department of Obstetrics and Gynecology, Inova Health System, Fairfax, VA US

**Keywords:** Medicaid Expansion, Affordable Care Act, Major Depressive Disorder, Women’s Health, Hospital Charges, Inpatient Care, Difference-in-Differences, Health Policy

## Abstract

**Objective:** To evaluate the effect of Medicaid expansion under the Affordable Care Act (ACA) on total inpatient hospital charges among women with MDD, comparing Maryland, an expansion state with an All-Payer Model, and Florida, a non-expansion state.

**Methods:** We conducted a retrospective cohort study using the Maryland State Inpatient Database and the Florida State Inpatient Database. The study population included women aged 18–64 years admitted with a primary diagnosis of MDD. The study period was stratified into pre-ACA (2007–2009) and post-ACA (2018–2020) eras. Difference-in-differences models with robust standard errors, complemented by inverse probability-weighted regression adjustment (IPWRA), were employed to estimate policy effects on hospital charges. Models adjusted for age, race/ethnicity, insurance type, discharge quarter, comorbidities, and neighborhood income quartile.

**Results:** A total of 122,963 hospitalizations were analyzed. Pre-ACA, baseline charges in Maryland averaged $7,123 (95% CI, 6,878–7,369). Following the ACA, Maryland experienced a moderated increase of $3,108 (95% CI, 2,752–3,465; p<0.001). In contrast, Florida’s charges remained significantly higher, exceeding Maryland by $5,136 pre-ACA and $11,413 post-ACA (p<0.001 for both). Difference-in-differences estimates confirmed that Medicaid expansion in Maryland mitigated cost escalation, producing a net relative reduction of $3,445 (95% CI, −3,886 to −3,004; p<0.001). Stratified analyses demonstrated the greatest financial protection among self-pay and Medicaid patients.

**Conclusion:** Medicaid expansion was associated with significantly lower inpatient charges among women with MDD in Maryland compared to non-expansion Florida, and may reduce financial burden, advance equity, and improve hospital resource sustainability.

## INTRODUCTION

Major depressive disorder (MDD) is one of the leading causes of disability worldwide^1,2^ and remains a major contributor to health care utilization and costs in the United States, particularly among women of reproductive age^3–5^. Beyond the profound individual and societal burden, inpatient psychiatric care accounts for a substantial proportion of expenditures related to MDD, and the steady rise in hospital charges has amplified concerns regarding affordability and equitable access to treatment^5–7^. Addressing the financial burden of psychiatric care and reducing disparities in access to essential services are therefore pressing public health priorities.

The passage of the Patient Protection and Affordable Care Act (ACA) in 2010 marked a landmark reform in U.S. health care, with provisions aimed at expanding insurance coverage, reducing the burden of uncompensated care, and improving access to essential services^8,9^. Among its most transformative elements was Medicaid expansion, which extended eligibility to millions of low-income adults^10,11^. Prior evidence suggests that Medicaid expansion has improved insurance continuity, access to mental health services, and reductions in unmet mental health needs^12–14^. These improvements are especially consequential for women with chronic mental health conditions such as MDD, for whom financial barriers often delay or limit care^15^.

Maryland and Florida offer a compelling natural experiment to assess ACA’s impact on hospital charges related to MDD. Maryland adopted Medicaid expansion in 2014 and operates under an All-Payer Model that regulates hospital payments across all payers, potentially moderating cost escalation^16,17^. In contrast, Florida did not adopt Medicaid expansion and continues to rely on traditional payment models^18^. This divergence in state policy creates an opportunity to disentangle the effects of Medicaid expansion from broader secular trends in hospital utilization and costs^19^.

The objective of this study was to evaluate whether Medicaid expansion under the ACA is associated with changes in inpatient hospital charges among women with MDD, comparing outcomes in Maryland (expansion state) with Florida (non-expansion state). By addressing this question, our study contributes new evidence at the intersection of health policy, women’s mental health, and hospital financing, providing insights into how large-scale policy reforms may influence financial protection and equity in care.

## Methodology

### Study Design and Data Source

We conducted a retrospective cohort study to evaluate the impact of Medicaid expansion under the Affordable Care Act (ACA) on inpatient hospital charges among women diagnosed with major depressive disorder (MDD). Data were obtained from the Healthcare Cost and Utilization Project (HCUP) State Inpatient Databases (SID)^20^, covering discharges between 2007 and 2020. The SID captures all inpatient encounters, providing a comprehensive representation of hospitalizations across states.

We selected Maryland as the Medicaid expansion state and Florida as the non-expansion comparator. Maryland adopted Medicaid expansion in January 2014 and is unique in its use of an All-Payer Model, which regulates hospital payments uniformly across payers, potentially moderating increases in hospital charges^16^. Florida, by contrast, did not expand Medicaid, making it an appropriate control to isolate the effect of expansion policies on financial outcomes.

### Study Population

The study population included women aged 18–64 years admitted with a primary or secondary diagnosis of major depressive disorder (MDD) (ICD-9 and ICD-10 codes). Patients ≥65 years were excluded due to near-universal Medicare eligibility, which would confound the impact of Medicaid expansion.

### Outcome Measures

The primary outcome for this study was total inpatient hospital charges (TOTCHG), a continuous measure that captures the financial burden associated with hospitalization. In the HCUP databases, TOTCHG represents the cleaned and edited total charges, while the unedited value is stored in TOTCHG_X. As part of HCUP’s standardized editing process, total charges are rounded to the nearest dollar, with zero charges set to missing. Charges deemed excessively low or high are recoded as inconsistent, with allowable ranges varying by year and database type. For example, in inpatient data, the permissible range was $25–$1.0 million from 1998– 2006, $100–$1.5 million from 2007–2010, $100–$5.0 million beginning in 2011, and $100–$10.0 million beginning in 2016. In general, professional fees and non-covered charges are excluded, though in rare instances they may be retained if state-specific data sources cannot separate them. Emergency department charges incurred prior to admission may also be included in TOTCHG, depending on payer billing requirements.

Secondary analyses examined stratified outcomes, including total charges by insurance type (Medicaid, private, Medicare, self-pay, other). This approach allowed us to evaluate whether the financial impact of Medicaid expansion varied across socioeconomic levels.

### Independent Variable

The primary independent variable was Medicaid expansion status, defined at the state level, with Maryland designated as the expansion state (coded as 1) and Florida as the non-expansion state (coded as 0). The analytic period was divided into two intervals: the pre-ACA period (2007– 2009) and the post-ACA period (2018–2020).

### Covariates

We adjusted for a broad range of demographic, socioeconomic, and clinical covariates to minimize confounding. Demographic variables included age (modeled as a continuous variable) and race/ethnicity (categorized as White, Black, Hispanic, Asian/Pacific Islander, Native American, or Other). Socioeconomic status was captured through median household income, stratified into quartiles (Q1–Q4). Insurance type was classified as Medicaid, private insurance, Medicare, self-pay, or other. Clinical factors comprised hypertension, obesity, diabetes, and smoking status, while hospitalization characteristics included discharge quarter to account for seasonality.

### Statistical Analysis

#### Descriptive Analysis

We first compared baseline characteristics across states and ACA periods using chi-square tests for categorical variables and t-tests/Wilcoxon rank-sum tests for continuous variables.

#### Inverse Probability Weighting (IPW)

To account for differences in patient characteristics across states and periods, we implemented inverse probability weighting. Propensity scores were estimated using logistic regression including age, race/ethnicity, insurance status, income quartile, and comorbidities. Each individual was weighted by the inverse of their probability of residing in an expansion versus non-expansion state, generating a pseudo-population balanced on observed covariates.

This method reduces confounding and allows for more robust causal inference. Weighted balance diagnostics were assessed using standardized mean differences.

#### Difference-in-Differences (DID)

The present study used a DID model to estimate the impact of Medicaid expansion on inpatient hospital charges among women presenting with major depressive disorder by comparing Maryland (treatment group, which implemented Medicaid expansion in 2014) to Florida (control group, which did not) during the pre-ACA and post-ACA periods. The variable ACA was set to 1 for the post-ACA period (2018–2020) and 0 for the pre-ACA period (2007–2009). We utilized an ordinary logistic regression with hospital charges as a continuous variable combined with an interaction between expansion status (Maryland vs Florida) and time-period (pre-ACA, 2007-2009 and post-ACA, 2018-2020).

The DID model is specified as: y=Xβ + β1·ACA+ β2·State + β12· (ACA X State).^21^

y represents the total inpatient hospital charges.

X includes selected covariates.

β1 captures the difference in outcomes between the pre- and post-ACA periods across both states.

β2 captures the baseline difference between Maryland and Florida.

β12 represents the effect of Medicaid expansion on total hospital charges.

The interaction term ACA × State (i.e., b12) estimates the difference in the change in total hospital charges between Maryland (the expansion state) and Florida (the non-expansion state) from the pre- to the post-ACA period. This term provides the key estimate of the impact of Medicaid expansion^21^.

#### Sensitivity Analysis

With a sensitivity analysis, we estimated models incorporating a three-way interaction between state Medicaid expansion status, ACA period (pre- vs. post-2014), and primary payer type. This approach allowed us to assess whether observed changes in total hospital charges were attributable primarily to Medicaid expansion rather than Maryland’s concurrent All-Payer Model. By stratifying the policy effect across payer categories (Medicaid, private insurance, self-pay, no charge, and other), we were able to test whether reductions in charges were concentrated among populations directly affected by expansion (Medicaid and self-pay) versus more uniformly distributed across all payer types, which would be more consistent with an All-Payer effect.

### Ethical Considerations

This study utilized de-identified HCUP SID data and was not considered human subjects research; therefore, Institutional Review Board approval and informed consent were not required.

## Results

### Patient Demographics and Clinical Characteristics

A total of 122,963 women hospitalized with major depressive disorder (MDD) were included, with 49,111 admissions in the pre-ACA period and 73,852 in the post-ACA period (Table 1). Florida consistently contributed the majority of hospitalizations, but Maryland’s representation allowed for comparison of an expansion versus non-expansion setting. Women admitted in the post-ACA era were slightly younger (mean age 36.4 ± 13.7 years in Maryland vs. 38.9 ± 14.2 years in Florida; p<0.01) (Table 1).

**Table 1.** Baseline Demographic and Clinical Characteristics of Patients in Maryland and Florida Pre- and Post-ACA Implementation.

| Variables<br>(N=122,963) | Pre-ACA (n=49,111 ) |  |  | Post-ACA (n=73,852) |  |  |
| --- | --- | --- | --- | --- | --- | --- |
|  | Florida 34,474 (70.2%) | Maryland<br>14,637<br>(29.8%) | P-value | Florida 61,509<br>(83.3%) | Maryland 12,343(16.7%) | p |
| Age, year, Mean ± SD | 41.9 ± 12.5 | 40.5 ± 12.1 | <0.01 | 38.9 ± 14.2 | 36.4 ± 13.7 | <0.01 |
| Race/Ethnicity |  |  | < 0.01 |  |  | < 0.01 |
| White | 25,750 (75.3%) | 4,241 (59.6%) |  | 39,343 (65.2%) | 6,590 (54.85) |  |
| Black | 3,768 (11.0%) | 2,411 (33.9%) |  | 9,762 (16.2%) | 4,016 (33.4%) |  |
| Hispanic | 3,313 (9.7%) | 160 (2.2%) |  | 9,536 (15.8%) | 775 (6.4%) |  |
| Asian or Pacific<br>Islander | 181 (0.5%) | 63 (0.9%) |  | 545 (0.9%) | 247 (2.0%) |  |
| Native American | 71 (0.2%) | - |  | 95 (0.2%) | 22 (0.2%) |  |
| Other | 1,096 (3.2%) | 238 (3.3%) |  | 1,059 (1.8%) | 379 (3.1%) |  |
| Median Household<br>Income |  |  | <0.01 |  |  | < 0.01 |
| Quartile 1 | 3,079 (25.8%) | 1,519 (21.8%) |  | 21,768 (37.0%) | 1,633 (13.4%) |  |
| Quartile 2 | 3,889 (32.6%) | 894 (12.8%) |  | 20,528 (34.9%) | 1,590 (13.1%) |  |
| Quartile 3 | 3,426 (28.7%) | 1,785 (25.6%) |  | 12,080 (20.5%) | 3,822 (31.4%) |  |
| Quartile 4 | 1,535 (12.9%) | 2,772 (39.8%) |  | 4,461 (7.6%) | 5,117 (42.1%) |  |
| Primary Pay |  |  | <0.01 |  |  | <0.01 |
| Medicare | 6,371 (18.5%) | 1,093 (14.8%) |  | 9,487 (15.4%) | 1,347 (10.9%) |  |
| Medicaid | 4,931 (14.3%) | 2,277 (30.9%) |  | 14,640 (23.8%) | 5,151 (41.9%) |  |
| Private Insurance | 16,566 (48.0%) | 3,023 (41.0%) |  | 22,274 (36.2%) | 4,729 (38.4%) |  |
| Self-Pay | 3,883 (11.3%) | 814 (11.0%) |  | 9,355 (15.2%) | 343 (2.8%) |  |
| No Charge | 833 (2.4%) | 35 (0.5%) |  | 3,191 (5.2%) | - |  |
| Other | 1,890 (5.5%) | 129 (1.7%) |  | 2,562 (4.2%) | 723 (5.9%) |  |
| HTN |  |  | < 0.01 |  |  | 0.031 |
| No | 26,396 (76.6%) | 10,516 (71.8%) |  | 48,432 (78.7%) | 9,611 (77.9%) |  |
| Yes | 8,078 (23.4%) | 4,121 (28.1%) |  | 13,077 (21.3%) | 2,732 (22.1%) |  |
| Obesity |  |  | < 0.01 |  |  | < 0.01 |
| No | 31,559 (91.5%) | 12,890 (88.1%) |  | 57,461 (93.4%) | 11,118 (90.1%) |  |
| Yes | 2,915 (8.5%) | 1,747 (11.9%) |  | 4,048 (6.6%) | 1,225 (9.9%) |  |
| DM |  |  | <0.01 |  |  | 0.325 |
| No | 31,220 (90.6%) | 12,912 (88.2%) |  | 57,731 (93.9%) | 11,556 (93.6%) |  |
| Yes | 3,254 (9.4%) | 1,725 (11.8%) |  | 3,778 (6.1%) | 787 (6.4%) |  |
| Smoking |  |  | <0.01 |  |  | <0.01 |
| No | 28,895 (83.8%) | 9,621 (65.7%) |  | 44,408 (72.2%) | 8,419 (68.2%) |  |
| Yes | 5,579 (16.2%) | 5,016 (34.3%) |  | 17,101 (27.8%) | 3,924 (31.8%) |  |
| Discharge Quarter |  |  | 0.155 |  |  | <0.01 |
| First | 8,126 (23.6%) | 3,590 (24.5%) |  | 15,337 (24.9%) | 3,206 (26.0%) |  |
| Second | 8,642 (25.1%) | 3,620 (24.7%) |  | 15,129 (24.6%) | 2,949 (23.9%) |  |
| Third | 8,998 (26.1%) | 3,760 (25.7%) |  | 15,787 (25.67%) | 2,967 (24.0%) |  |
| Fourth | 8,708 (25.3%) | 3,667 (25.0%) |  | 15,256 (24.8%) | 3,221 (26.1%) |  |

“-” symbols represent $\leq 10$ .

Racial and ethnic composition varied significantly across states and over time. Maryland had proportionally higher representation of Black women (33.9% pre-ACA; 33.4% post-ACA), while Florida was predominantly White and Hispanic (p<0.01). Socioeconomic differences were striking: over 40% of Maryland patients in the post-ACA era were from the highest income quartile compared with only 7.6% in Florida, whereas Florida had larger representation from the lowest quartiles (p<0.01) (Table 1).

Insurance coverage also shifted notably after the ACA. In Maryland, Medicaid coverage rose sharply (from 30.9% to 41.9%), while self-pay encounters fell markedly (from 11.0% to 2.8%; p<0.01). By contrast, Florida maintained higher rates of self-pay and “no charge” admissions. Comorbidities including hypertension, obesity, and smoking were consistently more prevalent in Maryland, reflecting a more complex hospitalized population (p<0.05 for each) (Table 1).

Together, these findings highlight important differences in baseline demographics, socioeconomic status, and payer mix across the two states, underscoring the need to adjust for patient- and system-level covariates in policy evaluation (Table 1).

### Overall Impact of Medicaid Expansion on Hospital Charges

Difference-in-differences models demonstrated that Medicaid expansion in Maryland was associated with a substantial reduction in total hospital charges compared with Florida (Table 2). Specifically, expansion corresponded to an adjusted decrease of $7,075 (95% CI, −$7,395 to −$6,755; p<0.001). Conversely, admission charges rose in the post-ACA period across both states, reflecting broader system-wide cost trends (+$6,080; 95% CI, 5,852 to 6,309; p<0.001). Importantly, the interaction term showed that the policy effect of expansion mitigated this upward pressure on costs, yielding a net reduction of $3,445 (95% CI, −$3,886 to −$3,004; p<0.001) for Maryland relative to Florida (Table 2).

**Table 2.** Adjusted Changes in Hospital Charges After Medicaid Expansion and ACA Period.

| Factor | Contrast (US\$) | Std. Err. | t | p value | 95% CI |
| --- | --- | --- | --- | --- | --- |
| EXPAND (Maryland vs base) | -7,075 | 163 | -43.31 | <0.001 | -7,395 to -6,755 |
| ACA (Post-ACA vs base) | 6,080 | 117 | 52.19 | <0.001 | 5,852 to 6,309 |
| EXPAND × ACA (Maryland × Post-ACA) | -3,445 | 225 | -15.30 | <0.001 | -3,886 to -3,004 |

Values represent difference-in-differences estimates of adjusted hospital charges associated with Medicaid expansion, ACA implementation, and their interaction. Estimates were obtained from linear regression models with robust standard errors. Results are expressed as mean changes (US dollars) with 95% confidence intervals (CIs).

These findings provide strong evidence that Medicaid expansion blunted the rising financial burden of psychiatric hospitalizations for women with MDD, contrasting with Florida where costs continued to escalate unchecked (Table 2).

### Sensitivity Analysis Using IPWRA

To strengthen causal inference, we conducted a sensitivity analysis using inverse probability-weighted regression adjustment (IPWRA) (Table 3). Results confirmed the robustness of our primary findings. Hospitalizations in Maryland post-ACA were associated with a mean increase of $3,108 compared with Maryland pre-ACA (p<0.001), reflecting secular trends. However, Florida’s costs were substantially higher, with pre-ACA charges exceeding Maryland by $5,136 and post-ACA charges surpassing Maryland by $11,413 (p<0.001 for both) (Table 3).

**Table 3.** Sensitivity Analysis Using IPWRA: Impact of Medicaid Expansion on Hospital Charges.

| Comparison | Coef. (US\$) | Std. Err. | z | p value | 95% CI |
| --- | --- | --- | --- | --- | --- |
| Maryland (Post-ACA) vs Maryland (Pre-ACA) | 3,108 | 182 | 17.07 | <0.001 | 2,752 to 3,465 |
| Florida (Pre-ACA) vs Maryland (Pre-ACA) | 5,136 | 409 | 12.56 | <0.001 | 4,334 to 5,937 |
| Florida (Post-ACA) vs Maryland (Pre-ACA) | 11,413 | 318 | 35.90 | <0.001 | 10,790 to 12,036 |
| POmean – Maryland (Pre-ACA) | 7,123 | 125 | 56.84 | <0.001 | 6,878 to 7,369 |

*Sensitivity analysis using inverse probability-weighted regression adjustment (IPWRA) to estimate the average treatment effect on the treated (ATET) for Medicaid expansion. Estimates are presented as mean changes in hospital charges (US dollars) with robust standard errors and 95% confidence intervals (CIs). POmean represents the predicted mean hospital charges for Maryland (Pre-ACA).*

The predicted outcome mean (POmean) for Maryland in the pre-ACA era was $7,123 (95% CI, 6,878 to 7,369), providing a clinically meaningful benchmark against which subsequent increases were measured. These findings reaffirm that expansion attenuated cost escalation, particularly when benchmarked against a high-cost, non-expansion environment such as Florida (Table 3).

### Impact Stratified by Insurance Type

Secondary analyses demonstrated clear heterogeneity in the impact of Medicaid expansion across payer categories (Table 4). The most pronounced benefits were observed among the populations most directly affected by coverage expansion. Self-pay patients experienced the largest reduction in hospital charges, with a mean decrease of $6,257 (95% CI, −$7,071 to −$5,442; p<0.001) (Figure 1). Medicaid-covered women also saw substantial savings, with charges reduced by $4,758 (95% CI, −$5,531 to −$3,986; p<0.001), underscoring the protective role of expanded coverage (Table 4).

**Figure 1.**
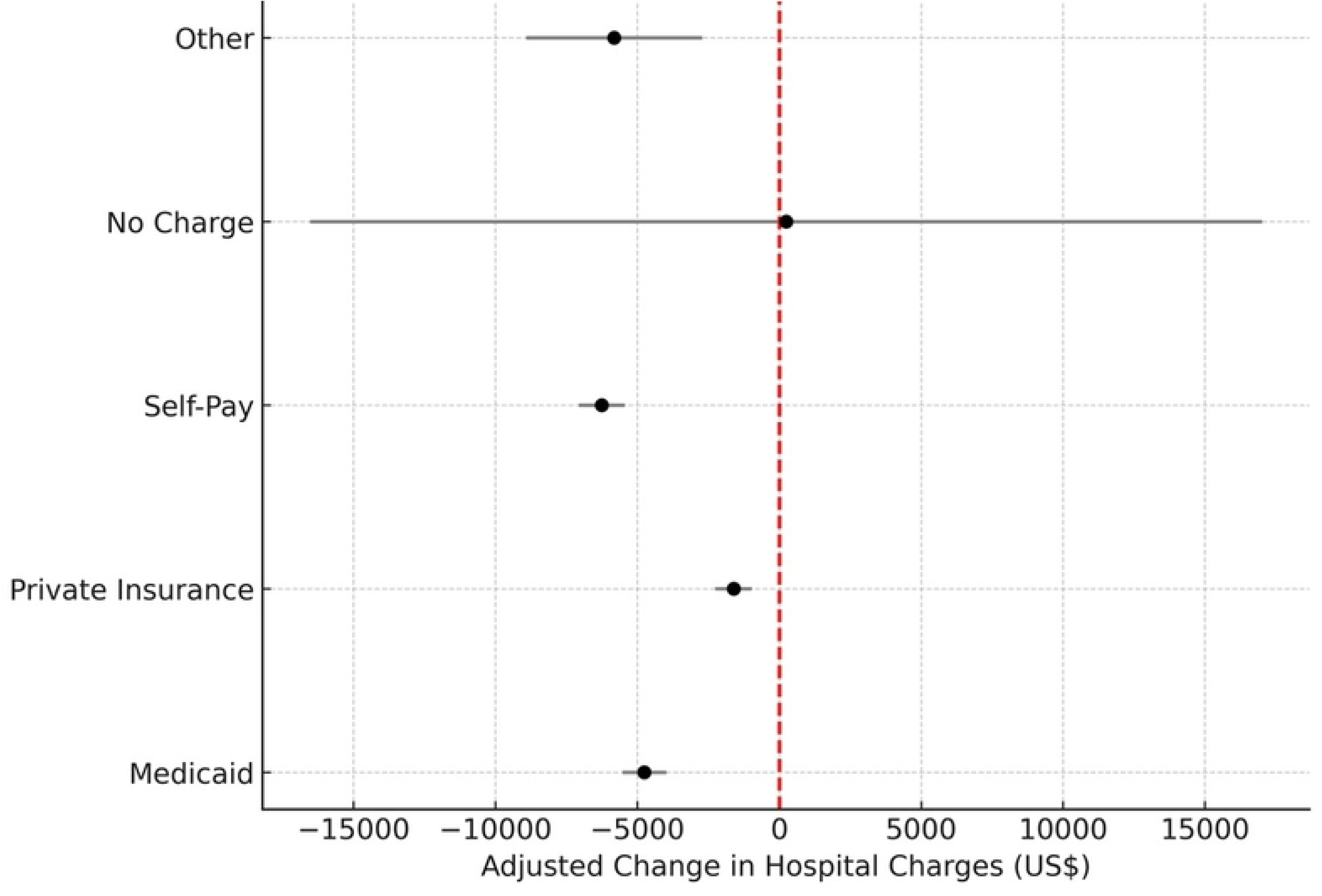
Adjusted Changes in Hospital Charges After Medicaid Expansion Stratified by Insurance Type (Excluding Medicare)

**Table 4.** Adjusted Changes in Hospital Charges After Medicaid Expansion, Stratified by Insurance Type.

| Insurance Type | Contrast (US\$) | Std. Err. | t | p value | 95% CI |
| --- | --- | --- | --- | --- | --- |
| Medicaid | -4,758 | 394 | -12.08 | <0.001 | -5,531 to -3,986 |
| Private Insurance | -1,608 | 329 | -4.89 | <0.001 | -2,253 to -964 |
| Self-Pay | -6,257 | 415 | -15.06 | <0.001 | -7,071 to -5,442 |
| No Charge | 243 | 8,553 | 0.03 | 0.977 | -16,522 to 17,008 |
| Other | -5,818 | 1,583 | -3.68 | <0.001 | -8,920 to -2,716 |

Values represent difference-in-differences estimates of adjusted hospital charges associated with Medicaid expansion, stratified by insurance type. Estimates were obtained from linear regression models with robust standard errors, incorporating a three-way interaction (*expansion* × *ACA period* × *insurance type*) and relevant covariates. Results are expressed as mean changes (US dollars) with 95% confidence intervals (CIs).

Smaller but statistically significant reductions were also noted among patients in the “other” insurance category (−$5,818; 95% CI, −$8,920 to −$2,716; p<0.001) and among privately insured women (−$1,608; 95% CI, −$2,253 to −$964; p<0.001). These findings suggest system-wide spillover effects, likely reflecting reduced uncompensated care burdens within hospitals. By contrast, no differences were observed in the “no charge” category (243; p=0.98), reinforcing that the financial benefits of Medicaid expansion were concentrated among patients whose coverage status directly determines hospital reimbursement (Table 4).

## Discussion

In this two-state comparative analysis of women hospitalized with major depressive disorders (MDD), we found that Medicaid expansion was associated with significant reductions in hospital charges, with the strongest effects observed among self-pay and Medicaid-covered patients. Using the Maryland State Inpatient Database (SID) and stratifying by pre-ACA (2007–2009) and post-ACA (2018–2020) periods, we demonstrated that Maryland’s adoption of Medicaid expansion in 2014 mitigated the upward trend in hospital costs that were observed in Florida. These findings reinforce the central role of health policy in shaping the financial burden of psychiatric care among women, a population disproportionately affected by economic and mental health challenges^4,22^.

Our results are consistent with prior studies demonstrating that Medicaid expansion reduces uncompensated care and hospital charges(23-26). For example, research by Nikpay et al. and Dranove et al. reported significant declines in uncompensated hospital costs following expansion, largely driven by reductions in self-pay admissions and increases in Medicaid coverage^27,28^. Similarly, studies examining other conditions such as acute cardiovascular events and surgical hospitalizations have shown lower total hospital charges or more favorable payer mixes in expansion states compared with non-expansion states^29–31^. By aligning with these prior findings, our analysis extends the evidence base into psychiatric care for women with MDD, a group less frequently examined in Medicaid policy research but at high risk of financial hardship due to recurrent hospitalizations^4,22^.

Several plausible mechanisms may explain the reductions in hospital charges observed in our study. First, Medicaid expansion reduces the proportion of uninsured patients, thereby shifting care from uncompensated or self-pay status to Medicaid reimbursement^29,30^. This transition not only lowers billed charges to patients but also stabilizes hospital finances, reducing cost-shifting pressures that can inflate charges for other payers^24,26,32^. Second, coverage expansion may encourage earlier and more consistent engagement with outpatient mental health services, potentially preventing disease exacerbation that leads to costlier inpatient stays^12,33^. Third, while Maryland’s All-Payer Model^16^ likely exerted some moderating influence by standardizing payment rates across payers, our sensitivity analysis confirmed that the observed reductions were not driven solely by this system. By incorporating a three-way interaction between expansion status, ACA period, and insurance type, we showed that cost reductions were most pronounced among Medicaid and self-pay patients the populations most directly affected by expansion while effects among privately insured women were smaller and consistent with spillover benefits^23^. Had the All-Payer Model been the primary mechanism, we would have expected more uniform reductions across all payer groups.

Our findings also suggest indirect “spillover” benefits to privately insured patients, echoing prior reports that expanded coverage can diffuse financial gains throughout the hospital system^23^. By reducing hospitals’ exposure to uncompensated care, Medicaid expansion may lessen the incentive to increase charges for privately insured populations, thereby creating a more equitable distribution of financial burden across payers^23,24^.

Taken together, our study highlights the potential of Medicaid expansion not only to protect the most vulnerable women but also to generate system-wide benefits through improved financial stability and more predictable reimbursement structures. These mechanisms underscore the broader value of expansion policies in reducing disparities in mental health care access and affordability.

### Policy Implications

The implications of these findings extend beyond hospital accounting and into the realm of public health and women’s rights. Women of reproductive age, who comprise a significant proportion of MDD hospitalizations, are particularly sensitive to gaps in insurance coverage. By reducing hospital charges, Medicaid expansion may help prevent catastrophic financial strain, improve continuity of mental health care, and indirectly support maternal and child health.

Importantly, our sensitivity analyses confirmed that these benefits were primarily attributable to Medicaid expansion, rather than to Maryland’s All-Payer Model. This distinction underscores expansion itself as the critical lever for change. Policymakers in non-expansion states, such as Florida, should consider the financial and equity benefits of expansion. Extending Medicaid coverage could alleviate systemic cost burdens, protect vulnerable women, and promote broader societal well-being by supporting access to care. For obstetrics and gynecology practitioners, these findings underscore the interconnectedness of mental health policy and women’s health outcomes, further emphasizing the need for integrated approaches to reproductive and behavioral health.

### Study Limitations

This study should be interpreted within the context of several limitations. First, our analysis relied on administrative data from the Maryland SID, which lacks granular clinical details such as symptom severity, pharmacotherapy use, and outpatient follow-up. Second, although we applied robust statistical methods, including sensitivity analyses with IPWRA, residual confounding cannot be excluded. Third, our pre- and post-ACA study windows (2007–2009 vs. 2018–2020) were intentionally chosen to capture stable baseline and post-implementation periods, but this design omits the transition years, which may have introduced additional dynamics in policy uptake. Fourth, while comparing Maryland and Florida provides a clear policy contrast, results may not be fully generalizable to other states with differing demographics, hospital systems, or reimbursement structures. Finally, Maryland’s unique All-Payer Model could independently influence hospital charges; however, our three-way interaction sensitivity analysis confirmed that reductions were concentrated among Medicaid and self-pay patients, suggesting that the observed effects were principally driven by Medicaid expansion rather than by all-payer rate setting alone.

## Conclusion

In summary, Medicaid expansion in Maryland significantly reduced hospital charges for women hospitalized with major depressive disorders, in contrast to Florida where costs continued to escalate. The largest benefits were observed among self-pay and Medicaid patients, with additional gains for privately insured populations. These findings highlight Medicaid expansion as a powerful lever for advancing health equity, protecting women from financial strain, and ensuring sustainable delivery of inpatient mental health care. More broadly, expansion policies represent not only an economic imperative but also a moral one, aligning financial systems with the principles of equity and compassionate care.

## Data Availability

Deidentified data underlying the results, along with analysis documentation, are available from the corresponding author upon reasonable request and contingent on required approvals and execution of a data use agreement.

## Acknowledgements

The authors acknowledge the support of the Clive O. Callender Outcomes Research Center at Howard University College of Medicine for this project.

## Author Contributions

O.A., M.F., F.A., G.K, S.K., O.O., R.O., C.E., A.O., M.M., and G.L., made substantial contributions to the conception and design of the study. O.A., F.O., and M.F. were involved in data acquisition, analysis, and interpretation. O.A., M.F., F.O., G.K., S.K., O.O., R.O., C.E., A.O., M.M., and G.L. contributed to drafting the manuscript. All authors critically reviewed the manuscript for important intellectual content and approved the final version. All authors have reviewed and approved the final version of the manuscript.

